# Locating cephalometric landmarks with multi-phase deep learning

**DOI:** 10.1101/2020.07.12.20150433

**Authors:** Soh Nishimoto, Kenichiro Kawai, Toshihiro Fujiwara, Hisako Ishise, Masao Kakibuchi

## Abstract

Cephalometric analysis has long been one of the most helpful approaches in evaluating cranio-maxillo-facial skeletal profile. To perform this, locating anatomical landmarks on an X-ray image is a crucial step, demanding time and expertise. An automated cephalogram analyzer, if developed, will be a great help for practitioners. Artificial intelligence, including machine learning is emerging these days. Deep learning is one of the most developing techniques in data science field. The authors attempted to enhance the accuracy of an automated landmark predicting system utilizing multi-phase deep learning and voting. To guarantee objectivity, an open-to-the-public dataset, cephalometric images accompanied with coordinate values of 19 landmarks, were used. A regressional system was developed, consisted with convolutional neural networks of three phases. First phase network was to determine approximate position of each landmark, inputting whole area of compressed original images. Five secondary networks were to narrow down the area, based on the first phase prediction. Third phase networks were trained by small areas around respective landmarks, with original resolution. Third phase prediction with voting was done inputting 81 shifted areas. Successful detection rates improved as the phase advances. Voting in third phase improved successful detection rate. In comparison with previously reported benchmarks, using the same dataset, proposed system marked better results. Within the physical limitation of memory and computation, multi-phase deep learning may be a solution to deal with large images.

## Introduction

Cephalometry was first introduced by Broadbent in1931^1^. X-ray image was taken, stabilizing the examinee’s head in a standard position. It has been and still is one of the most helpful modalities in evaluating cranio-maxillo-facial relations. In analyzing process, plotting landmarks on the X-ray image is required. This procedure requires time and expertise. Machine learning is one of the most developing fields in data science. Artificial neural network^2,3^ is a type of approach in machine learning. Optimizing artificial neural network with many layers is called deep learning^4^. Convolution neural network^5^,^6^ is a type of deep learning method with one or more convolutional layers. It has been attracting attention in image recognition. In medical field, cognition of dermatological images^7-9^, X-ray images^10^, optical funduscopic images^11^ and coherence tomography^12^. Convolution neural network can also be used for regression^13,14^.

In a previous report, we built an automated landmark prediction system, based on a deep learning convolutional neural network^15^. Cephalograms were gathered through the internet and landmarks were plotted by ourselves. Although it seemed to be a good result, objectivity was not guaranteed. In this study, an open-to-the-public dataset was used to evaluate our system objectively. 2014 IEEE 11th International Symposium on Biomedical Imaging (ISBI)^16^, and 2015 IEEE 12th ISBI^17^ was done and cephalograms with coordinate value for each landmark is released to the public (http://www-o.ntust.edu.tw/~cweiwang/ISBI2015/). In one staged deep learning, original high-resolution (2400 x 1935) images have to be resized into low-resolution ones, to reduce computation volume. Precise information may be lost in this process. To increase prediction accuracy, multi-phased deep learning was conducted.

## Materials and Methods

### Personal computer

All procedures were done on a desk-top personal computer: CPU (Central Processing Unit): AMD Ryzen 7 2700X 3.70GHz (Advanced Micro Systems, Sunnyvale, CA, USA), memory: 64.0GB, GPU: GeForce RTX2080 8.0GB ((nVIDIA, Santa Clara, CA, USA), Windows 10 pro (Microsoft Corporations, Redmond, WA, USA). Python 3.6 (Python Software Foundation, DE USA): a programing language, was used under Anaconda (FedoraProject. http://fedoraproject.org/wiki/Anaconda#Anaconda_Team_Emeritus) as an installing system, and Spyder 3.6 16 as an integrated development environment. Keras (https://keras.io/): the deep learning library, written in Python was run on TensorFlow (Google, Mountain View, CA, USA). GPU computation was employed through CUDA (nVIDIA). OpenCV3.1.0 libraries (https://docs.opencv.org/3.1.0/) were used in image processing.

### Datasets

Lateral cephalograms with coordinate values of 19 landmarks each were downloaded from https://figshare.com/s/37ec464af8e81ae6ebbf. They were the ones published and used in 2015 IEEE 12th ISBI Challenge 1. Original resolutions of them were 1935 pixels for horizontal (x) and 2400 pixels for longitudinal (y), representing 0.1 mm/pixel. Coordinate values of the landmarks are provided in a text file for an image. There provided two sets of them in different folders, named “400_junior” and “400_senior”. The coordinate values of all images in a folder were gathered into a table and saved as a csv formatted file. To be in conformity with previous studies, average coordinate values of the two sets were used as ground truth (Fig 1).

**Fig 1.**
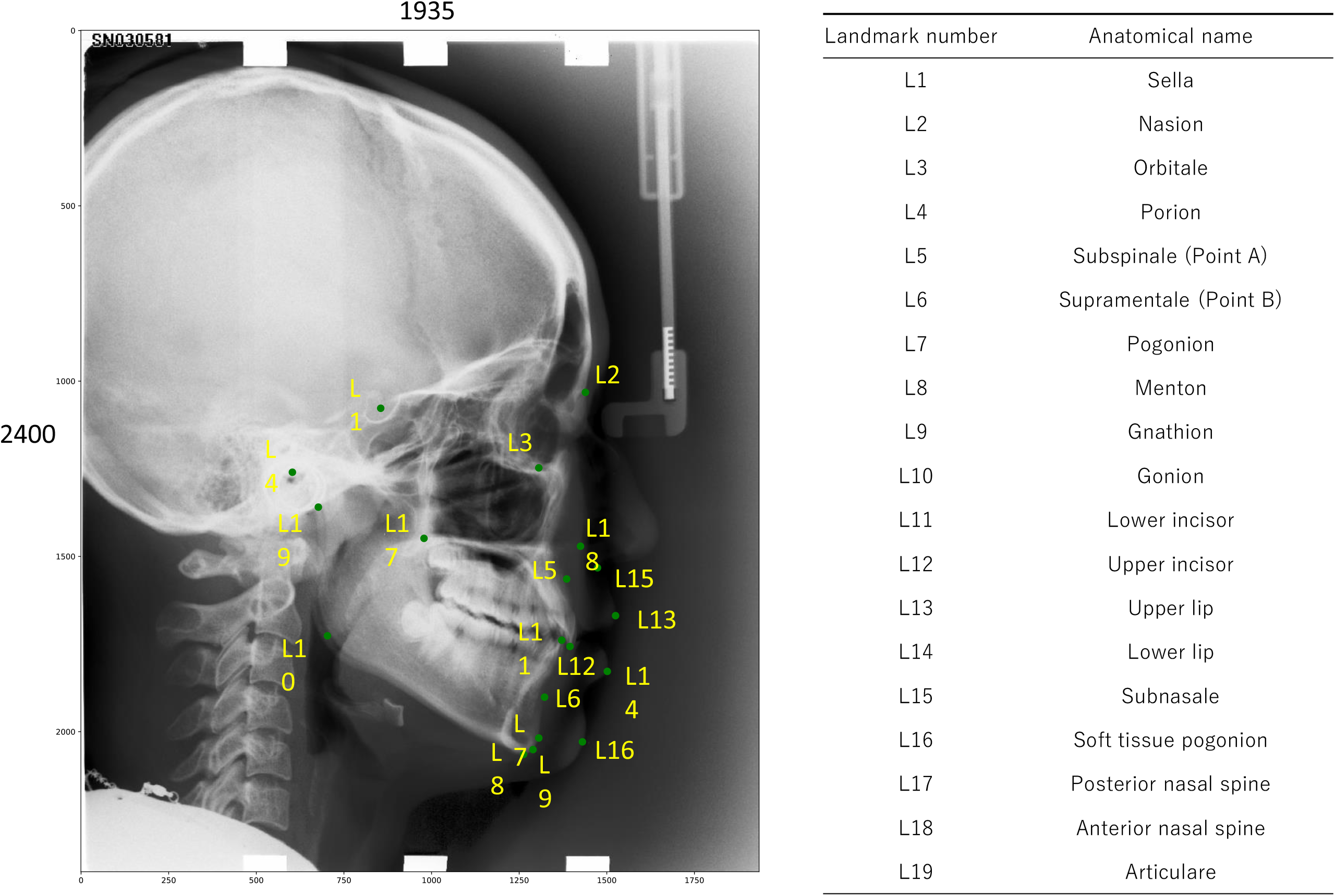
Anatomical landmarks plotted on a cephalogram (TrainingData/009.bmp). Coordinate values of them were set as the average of the two provided sets.

### Multi-phase deep learning

#### First phase deep learning

Images for training, counting 150, were loaded in grayscale and downsized to 387 x 480 pixels.

Data augmentation was done to increase training images (Fig 2). Gamma values of the images was changed using OpenCV program. Shifting and rotation of the images were performed, with coordinate values of the landmarks correspondingly. Water-droplet-like distortion of the images^18^ were also done with corresponding coordinate values computation. When any landmark placed out of range, the data was omitted. Yield of 18288 images was obtained.

**Fig 2.**
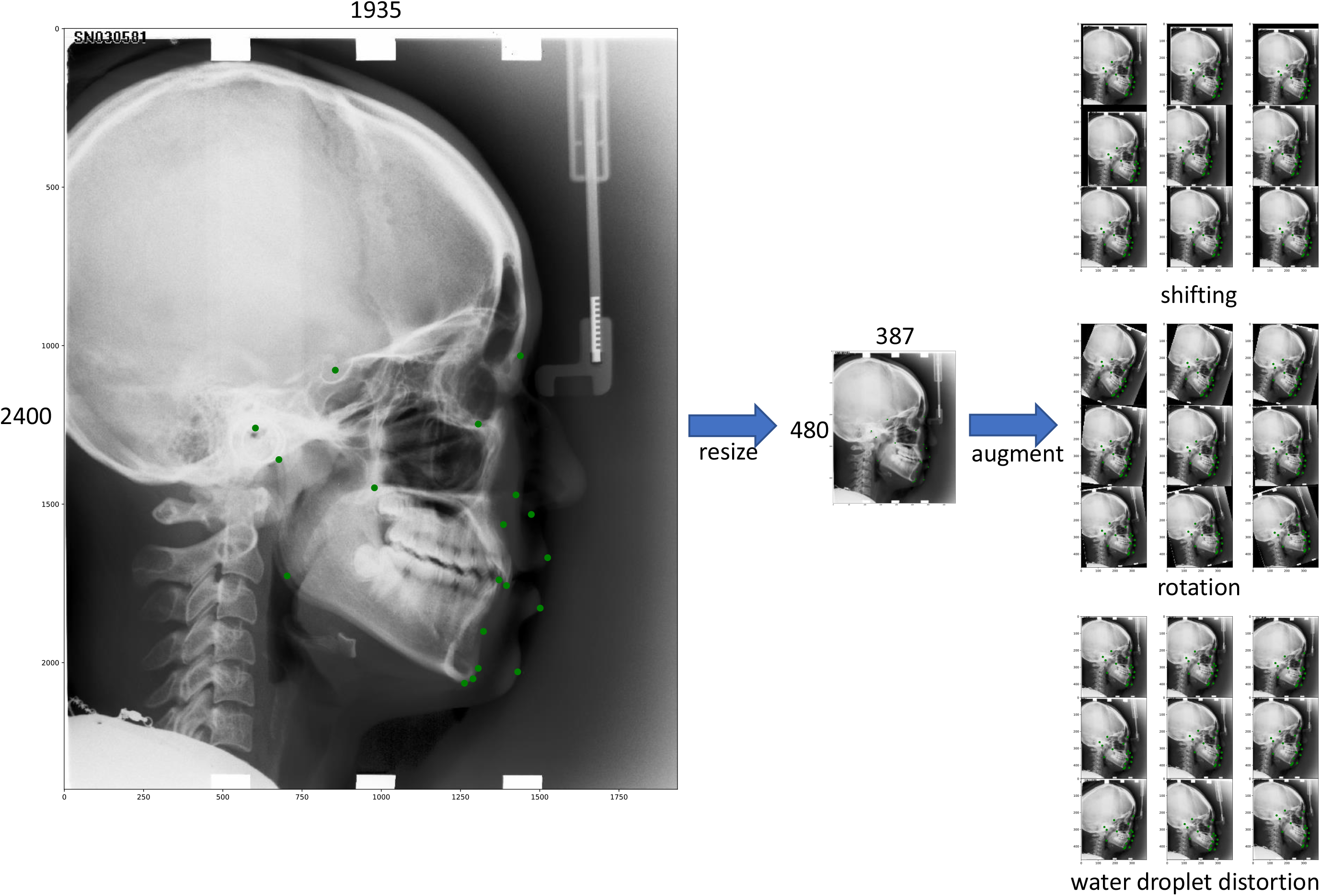
For the first phase deep learning, training images, originally 1935 x 2400 pixels, were downsized to 387 x 480 pixels. For data augmentation, shifting, rotation and water-droplet-like distortion of the images were performed, with the change of landmarks coordinate values correspondingly. Shown is an example (TrainingData/009.bmp).

Based on Keras with TensorFlow backend on Python, a regression neural network with 4 convolution layers, followed by 4 dense layers, which was used in our previous report^15^ (Fig 3), was constructed, with input of 387 x 480 and output of 38 values (x, y each for 19 landmarks). Stochastic gradient descent was used as the optimizer. The Leaky-ReLu was used as activating function. Epochs of 800 were done, trained with previously described 18288 images and corresponding coordinate values. The model and trained weights were saved in json and h5 formatted files, to use in further prediction.

**Fig 3.**
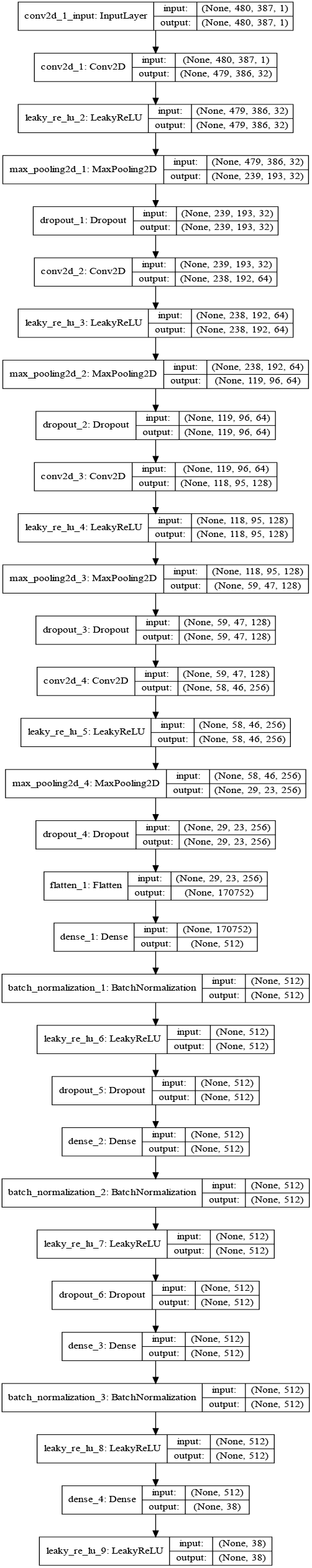
The convolutional neural network model, used in the first phase deep learning. Ii is consisted of 4 convolution layers and 4 dense layers. Basically the same model was used for the second and third phase deep learning, except for the input and output shapes.

#### Second phase deep learning

Landmarks were divided into five groups: upper posterior (L1: Sella, L4: Porion, L19: Articulare), upper frontal (L2: Nasion, L3: Orbitle), upper jaw (L5: Point A, L12: Upper incisor, L13: Upper lip, L15: Subnasalis, L18: Anterior nasal spine), lower jaw (L6: Point B, L7: Pogonion, L8: Menton, L9: Gnathion, L11: Lower incisor, L14: Lower lip, L16: Soft tissue pogonion) and lower posterior (L10: Gonion, L17: Posterior nasal spine). For each group, the center coordinate values of the landmarks were calculated. Sub-areas of 600 x 600 pixels were cropped from the original image with the previously calculated centers (Fig 4). Areas with the centers, shifted −200 to 200 pixels, stepping 40 pixels, were also cropped. The relative coordinate values of the landmarks in the cropped images were listed. The images were downsized to 200 x 200 pixels. Data augmentation was done by enlarging and making small, accompanied by coordinate values changes. Main structure of the network model was the same as the one used for the first phase deep learning. Input was 200 x 200 and output numbers were varied respectively. Training of the models were done with the epochs of 500. The models and trained weights were saved in json and h5 formatted files.

**Fig 4.**
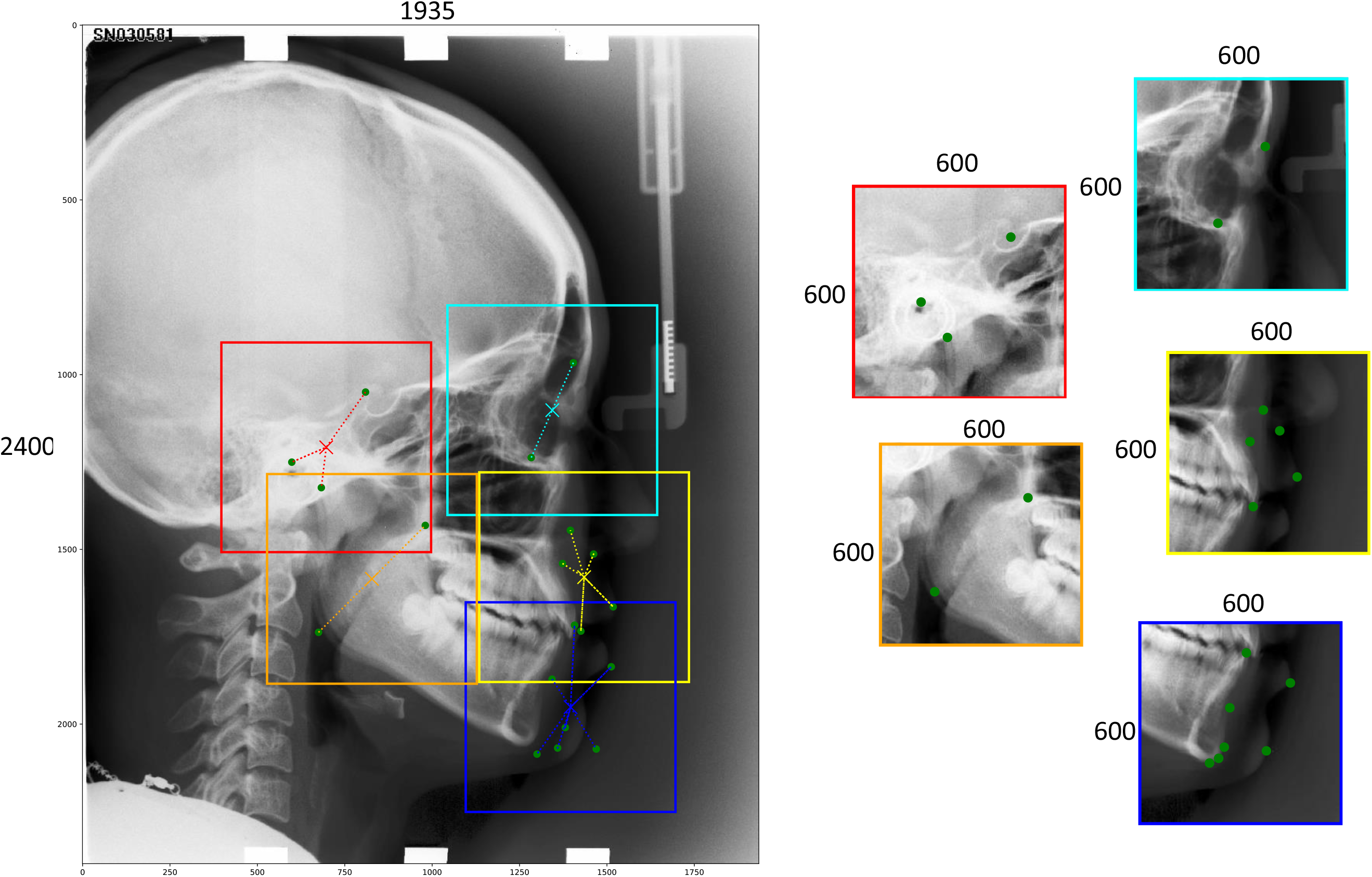
Landmarks were divided into five groups. Midpoint of each group was set as the center for 600 x 600 pixels sub-areas. The Sub-areas were cropped for the training images for the second phase deep learning. Shown is an example (TrainingData/009.bmp).

#### Third phase deep learning

Setting the landmarks as the centers, 200 x 200 pixels sized image were cropped (Fig 5). Areas, shifted −50 to 50 pixels stepping 10 pixels, were cropped and coordinate values of the landmark were calculated as the relative values in the image. The images were processed with CLAHE (contrast limited adaptive histogram equalization)^19^ using OpenCV. Data augmentation was done as the second phase deep learning. Basic structure of the neural model was the same as the first and second deep learning, except that the input was 150 x 150 and the output was two (x, y). The model was trained with the epoch of 500 and saved.

**Fig 5.**
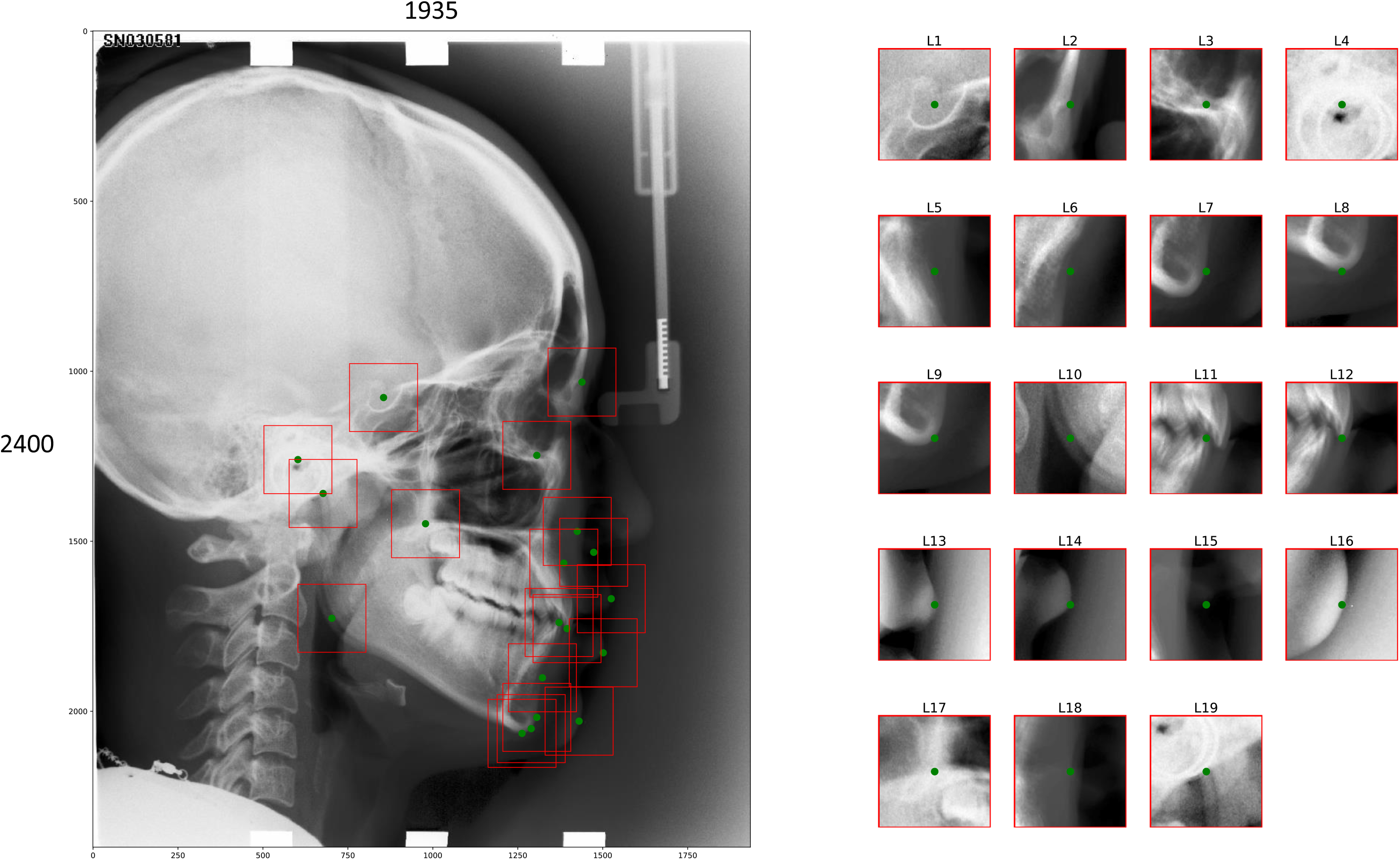
200 x 200 pixels sized areas were cropped, setting landmarks as the centers. They were used to train the third phase convolutional neural networks. Shown is an example (TrainingData/009.bmp).

### Landmark prediction

Flow chart for landmark prediction and evaluation is shown in Fig 6.

**Fig 6.**
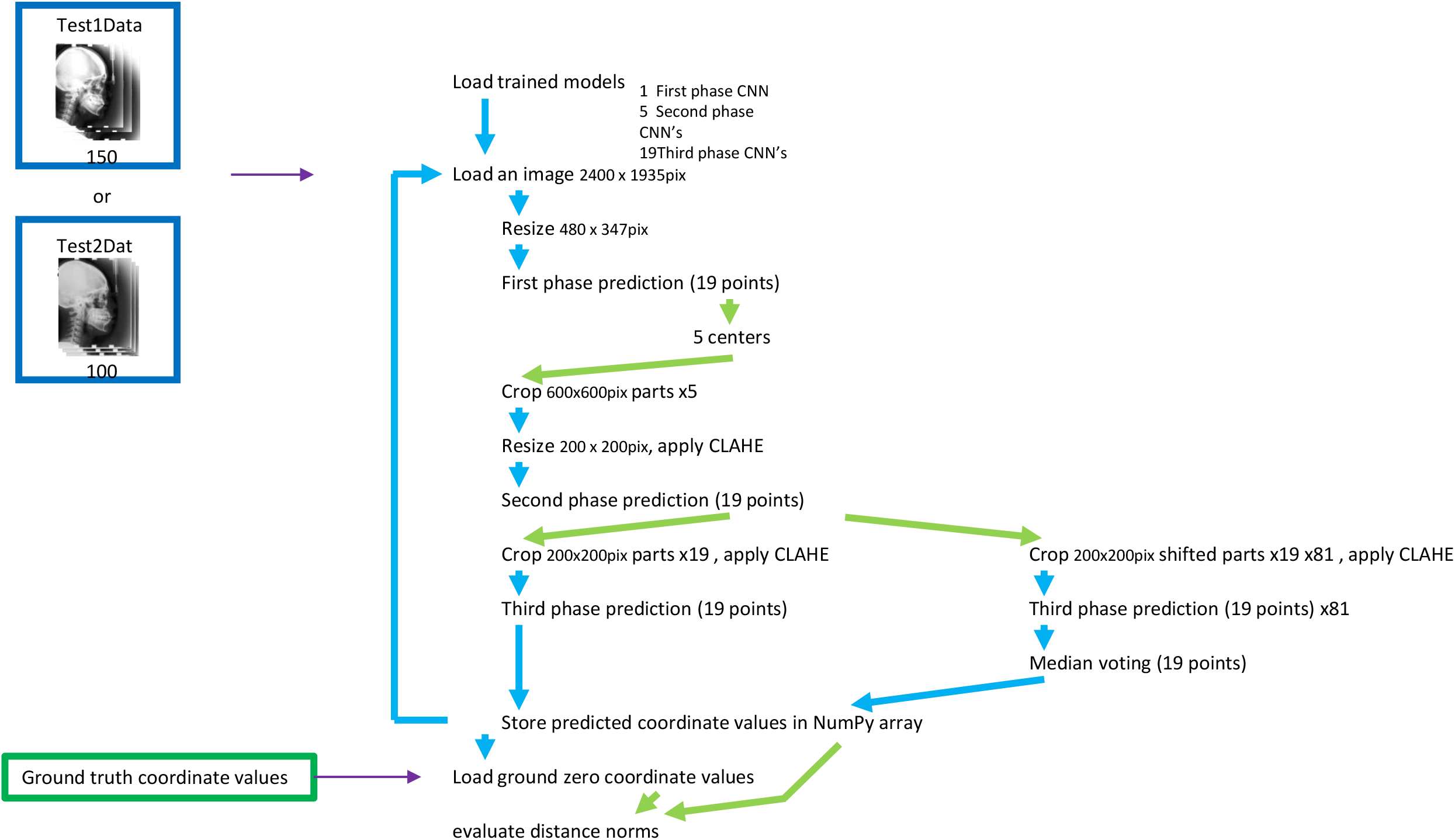
Flow of landmark prediction and evaluation. Coordinate values of each landmark were predicted through three trained convolutional neural networks.

#### First phase prediction

Testing images, which were not used for training, were loaded in grayscale, downsized to 387 x 480 pixels and fed to the first phase trained model. 38 values (x, y for each19 points) for every image were predicted.

#### Second phase prediction

Based on the first phase coarse prediction, 600 x 600 pixel-sized sub-areas were cropped from the original testing images. They were downsized to 200 x 200 pixels, CLAHE processed and fed to the second phase trained models.

#### Third phase prediction

Setting coordinate values, obtained by the second phase prediction, as the center points, 19 areas with 150 x 150 pixels were cropped from each original testing images. The images were CLAHE processed, fed to the trained models and coordinate values for each landmark were predicted.

#### Third phase prediction with voting

Eighty-one areas, shifted the second phase predicted centers −20 to 20 pixels stepping 5 pixels, were cropped from each original testing images. The images were CLAHE processed, fed to the trained models and coordinate values for each landmark were predicted. Median of the predicted 81 coordinated values was adopted as the prediction.

### Evaluation of the predicted coordinate values

Distance between a point, predicted by neural network P: (px, py) and corresponding ground truth point O: (ox, oy) was calculated as norm of vector OP: (px - ox, py - oy). One pixel-length was evaluated as 0.1 mm. Successful detection rate, defined as the percentage of landmarks predicted within 2.0 mm, 2.5 mm, 3.0 mm and 4.0 mm, was calculated. Within a series of procedures, prediction of the testing images, which were not used in the training stage, were done one by one, followed by evaluation by NumPy computation.

## Results

### Machine learning

#### First phase deep learning

Learning for 800 epochs for the first phase with 18288 training images, took 57 hours and 47 minutes.

#### Second phase deep learning

Average testing image number used for each second phase was 14578. Average duration time for learning 500 epochs was 3 hours and 35 minutes.

#### Third phase deep learning

Number of images, used for the third phase deep learning, was 47248 respectively for each landmark. It took 1 hour 55 minutes and 5 seconds in average for 100 epochs training.

### Landmark coordinate value prediction

To load modules of TesorFlow, Keras, OpenCV and so on, it took 1.45 seconds in our setting. To load neural networks and weights, it took 39.16 seconds. For 150 datasets of Test1Dataset, loading images and coordinate value predicting with three phase networks required 42.74 seconds. Computing distances from ground truth landmarks took 0.29 seconds. For Test2Dataset, it took 18.47 seconds for coordinate value prediction and 0.22 seconds for evaluation. With voting, prediction took 15 minutes and 55 minutes for Test1Dataset and 6 minutes and 54 seconds for Test2Dataset, Successful detection rates for each three phases assessing Test1Dataset and Test2Dataset is shown respectively in Fig 7 and 8. Landmark by landmark prediction errors (distances from ground truth coordinate values) are shown in Fig 9.

**Fig 7.**
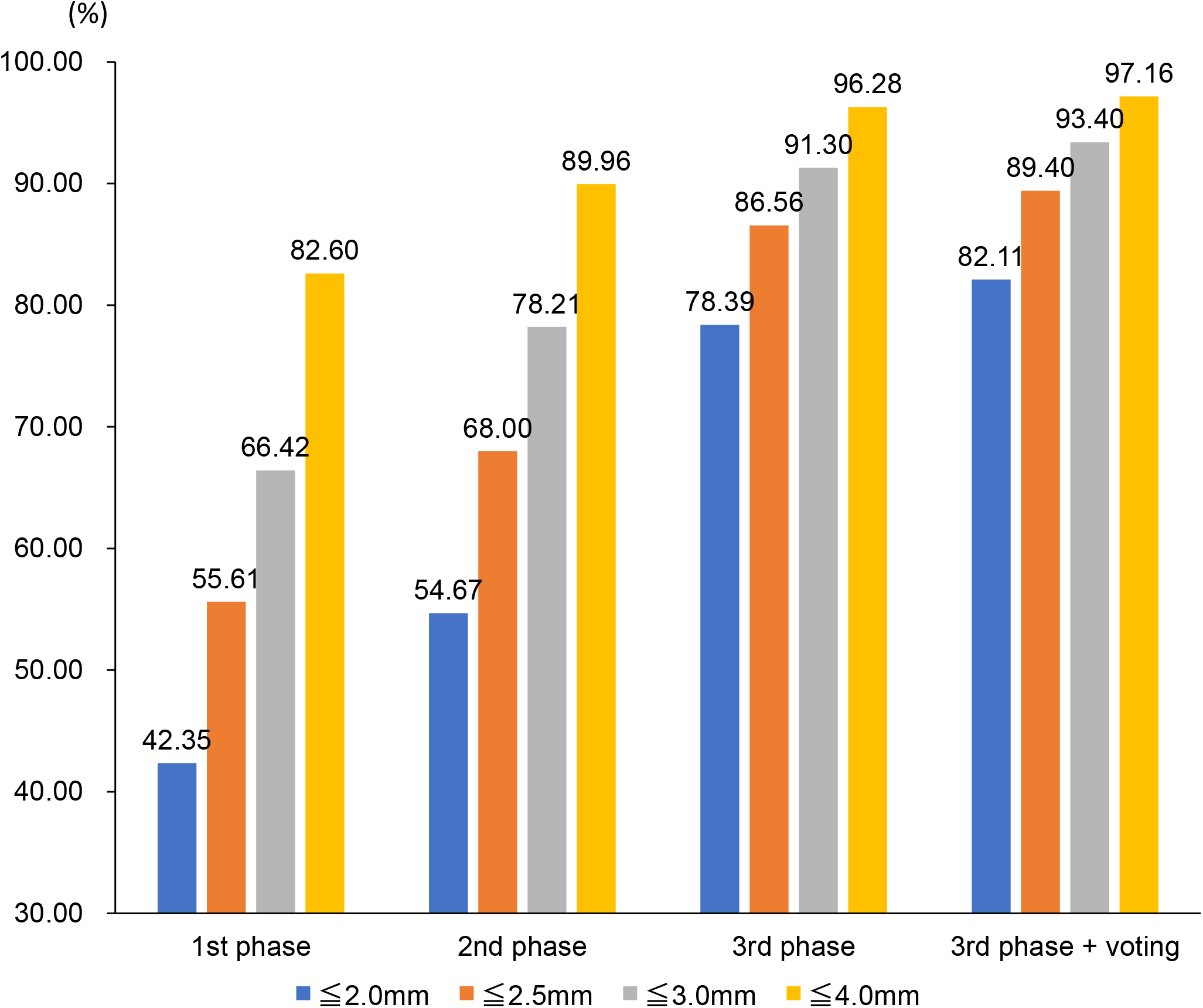
Successful detection rates for 150 images of Test1Dataset. The rates increased as the phase advances. Third phase with voting marked the best result.

**Fig 8.**
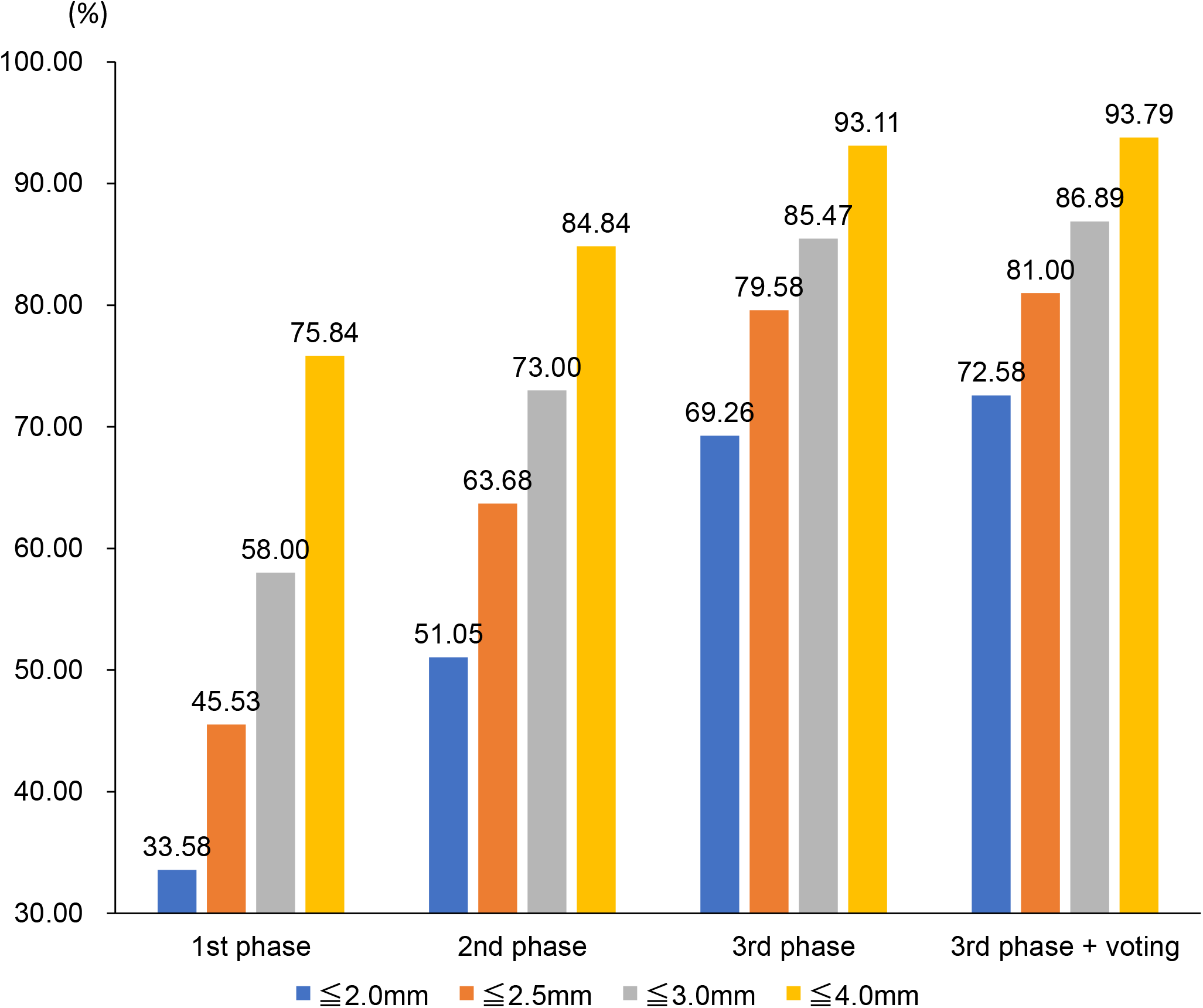
Successful detection rates for 100 images of Test2Dataset. The rates increased as the phase advances. Third phase with voting marked the best result.

**Fig 9.**
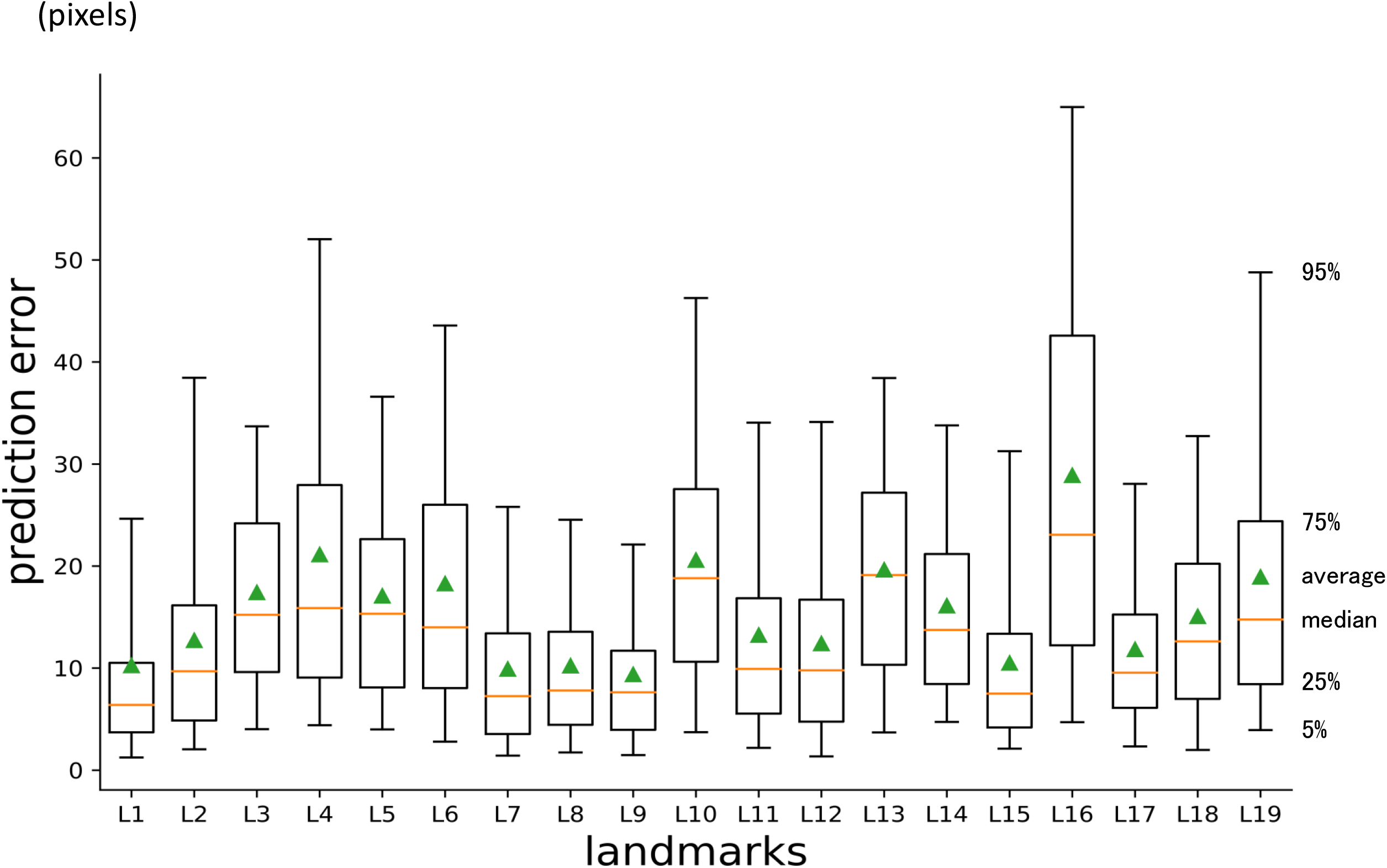
Box plotting of prediction errors (distance between predicted point and ground truth point) for each landmark, testing 250 images (Test1Dataset and Test2Dataset). Whisker ranges 5 to 95 percntile. Box ranges 25 to 75 percentile. Center line indicates the median. Triangle indicates the average.

## Discussion

Our concept was to emulate the way in finding a place, when the address is provided. First, we open a map of the country. Next, we try to find the state and city in a larger scaled map. Then, open a much larger scaled map to find the street and number. You cannot precisely point the place in a low-scaled map. And, you cannot orientate yourself in a large-scaled map from the first.

First phase prediction was done by a network, which evaluate whole area of an image. The original image to evaluate was 1935 x 2400 pixels. They were downsized to 387 x 480 pixels because of computation limitation. Second phase prediction was done in smaller areas. Interested area of 600 x 600 pixels size, based on the coarse prediction by the first deep learning model, was cropped and downsized to 200 x 200 pixels. Third phase prediction was done in cropped images from the original one, with the size of 200 x 200 pixels, based on the coordinate values of second prediction. Prior to this, we tried with 150 x150 pixels areas (data not shown). Better results were obtained with 200 x 200 pixels. As shown in Fig 7 and 8, Successful detection rates improved as the phase advances. Within the physical limitation of memory and computation, multi-phase deep learning may be a solution to deal with large images. CLAHE processing before second and third phase prediction slightly improved accuracy (comparison with or without is not shown). Voting in last phase improved successful detection rate.

Comparing with previously reported benchmarks (Ibragimov and Lindner: the two best in ISBI2015^17^, Arik^20^: a method using convolutional neural network), successful detection rate by proposed procedures marked better results (Fig 10,11).

**Fig 10.**
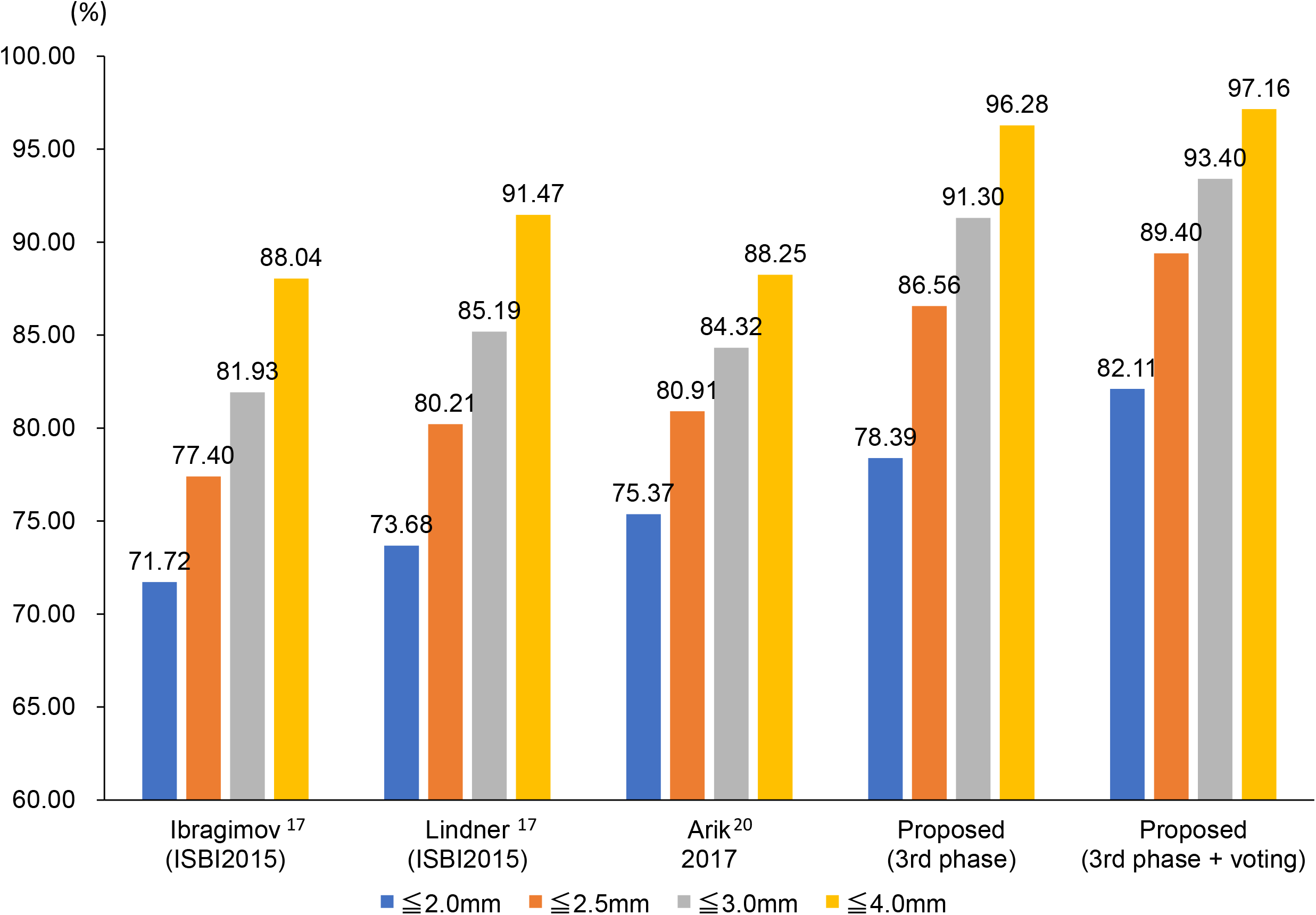
Comparison of previously reported benchmarks with proposed procedures, in successful detection rates for Test1Dataset.

**Fig 11.**
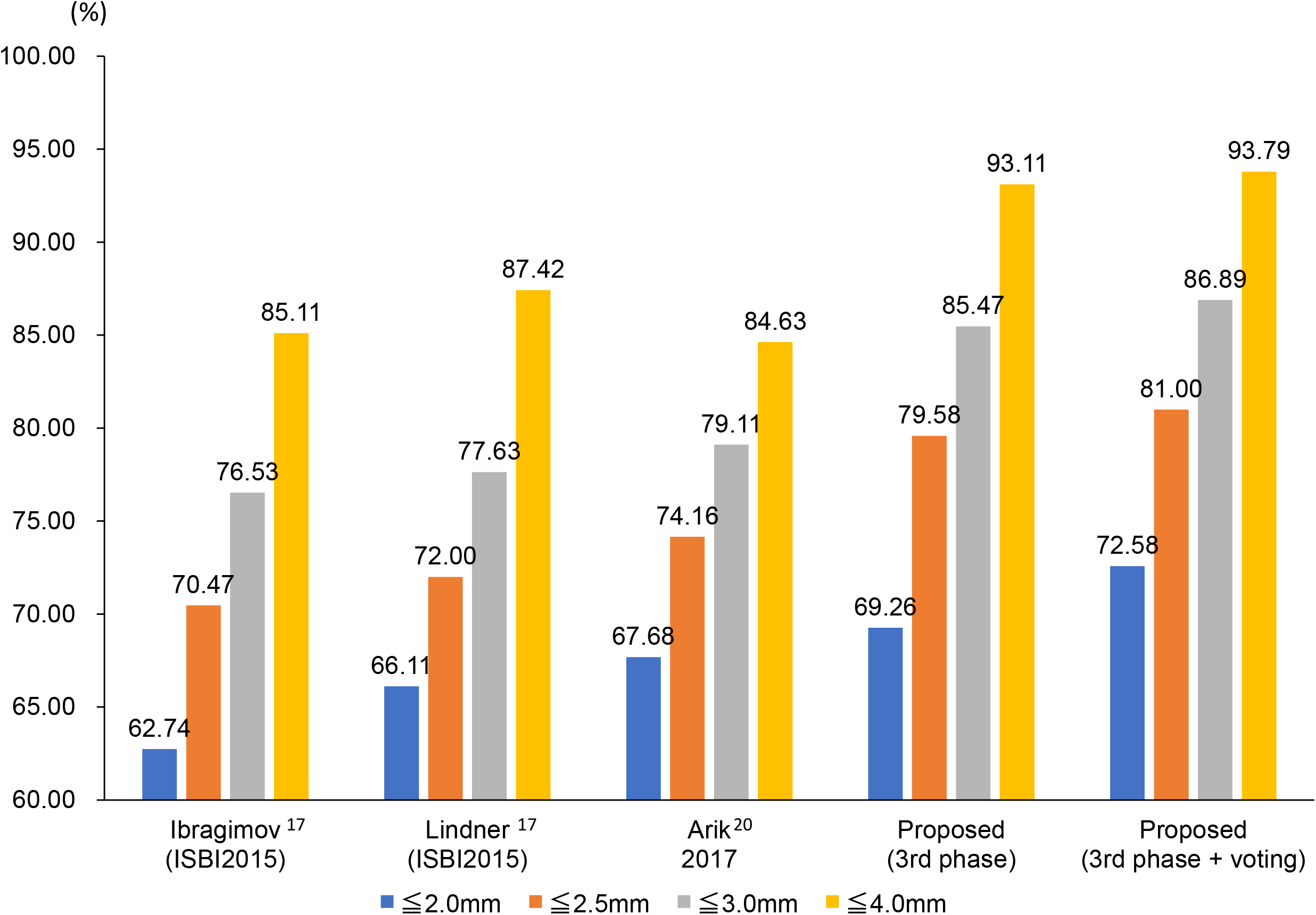
Comparison of previously reported benchmarks with proposed procedures, in successful detection rates for Test2Dataset.

It is well known that accuracy of machine learning prediction highly be dependent on the number of training data. Though, there are many cases that the number of raw data is limited. Data augmentation technique helped making up for the data shortage. It markedly improved our accuracy (data not shown).

It has been pointed that manual plotting can be varied by plotters^16,21^ or time even by the same plotter^16,22^. Ground truth coordinate values may differ from one plotter to the other. How much error can be clinically and practically allowed is hard to standardize. In the datasets, we used in this study, there provided two sets of coordinate values (“400_junior” and “400_senior”), plotted by two different practitioners. Comparing 7600 landmark points coordinate values in 400 images, there existed 2.15 mm discrepancy in average with 2.22 mm standard deviation, 21.7mm as maximum. Landmark by landmark discrepancies of them in the 150 training images are shown in Fig 12. Largest discrepancy was seen on L16: Soft tissue pogonion, which is not always easy to plot even by experienced practitioners.

**Fig 12.**
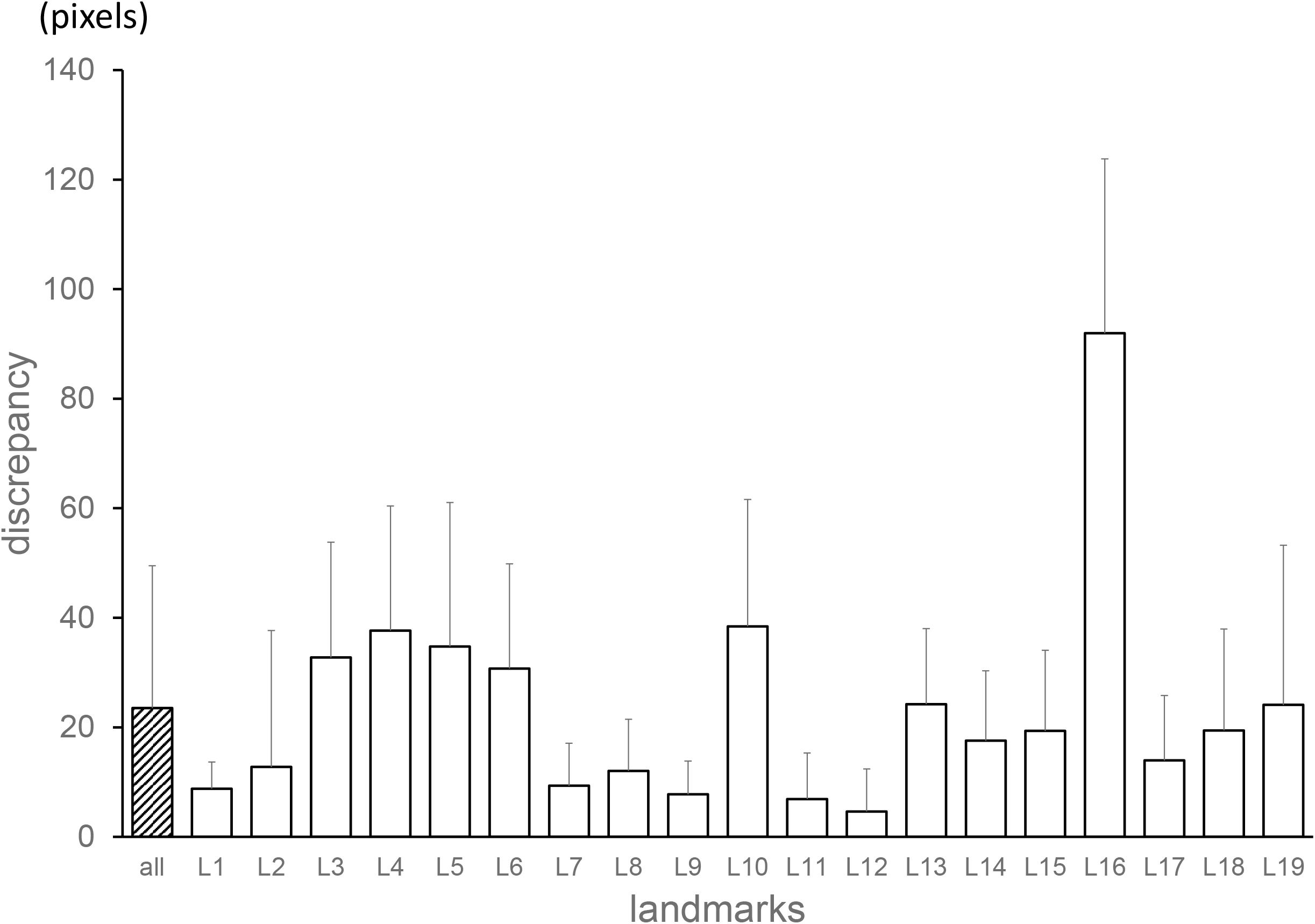
Discrepancies between the two provided coordinate values (in folders, named “400_junior” and “400_senior”) for training images. 2850 points for all and 150 points for each landmark. Average + standard deviation.

In this study, we used the mean value of them to be in conformance with former studies. This averaging process may have led to that the point is not be on the edge of structure, which could complicate the predictor.

Landmarks were plotted in Fig 13, with average discrepancies between the two provided coordinate values for the training datasets as horizontal axis and average prediction errors for the testing datasets in our best scored system as vertical axis. Very high correlation was seen between them. It implies that difficulty in plotting the landmarks influenced the prediction accuracy.

**Fig 13.**
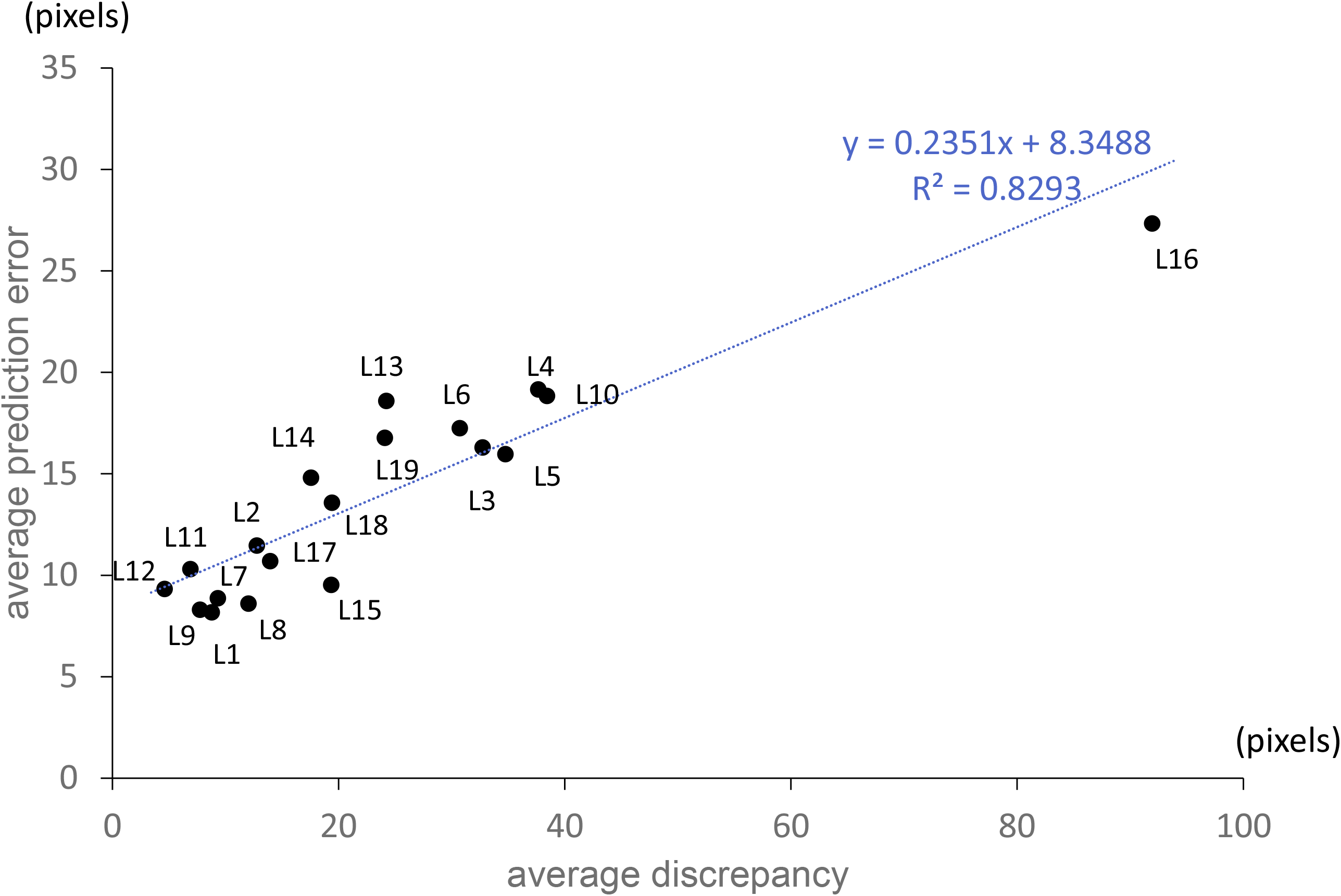
Landmark by landmark plotting. Horizontal axis: average discrepancies between the two provided coordinate values (“400_junior” and “400_senior”) in 150 training datasets. Vertical axis: average prediction errors for the 250 testing datasets by 3rd phase + voting system. With linear regression analysis, high correlation was observed.

In supervised learning and prediction, high prediction implies that ground truth values were consistently rated in training and testing datasets. If the training data plot is not clinically “correct”, the predicted value will not be “correct”. High quality coordinate values in training datasets are essential.

## Conclusion

We constructed an automated cephalogram landmark plotting system, utilizing multi-phase convolutional neural networks. The system was consisted of three phases. Each phase was trained respectively. The system significantly increased accuracy comparing with a single-phased model. As there always physical limitation in computation, multi-phase deep learning may be a solution to deal with large images.

## Data Availability

None

## Acknowledgement

There is no financial conflict of interest.

